# Serologic Surveillance and Phylogenetic Analysis of SARS-CoV-2 Infection in Hospital Health Care Workers

**DOI:** 10.1101/2021.01.10.21249440

**Authors:** Jonne J. Sikkens, David T.P. Buis, Edgar J.G. Peters, Mireille Dekker, Michiel Schinkel, Tom. D.Y. Reijnders, Alex. R. Schuurman, Justin de Brabander, Ayesha H.A. Lavell, Jaap. J. Maas, Jelle Koopsen, Alvin X. Han, Colin A. Russell, Janke Schinkel, Marcel Jonges, Sebastien P.F. Matamoros, Suzanne Jurriaans, Rosa van Mansfeld, W. Joost Wiersinga, Yvo M. Smulders, Menno D. de Jong, Marije K. Bomers

**Affiliations:** Amsterdam UMC, Vrije Universiteit Amsterdam, Department of Internal Medicine, Amsterdam Infection and Immunity Institute, De Boelelaan 1117, 1081 HV, Amsterdam, Netherlands; Amsterdam UMC, Vrije Universiteit Amsterdam, Department of Internal Medicine, Division of Infectious Diseases, Amsterdam Infection and Immunity Institute, De Boelelaan 1117, 1081 HV, Amsterdam, Netherlands; Amsterdam UMC, Vrije Universiteit Amsterdam, Department of Medical Microbiology & Infection Prevention, Amsterdam Infection and Immunity Institute, De Boelelaan 1117, 1081 HV, Amsterdam, Netherlands; Amsterdam UMC, University of Amsterdam, Center for Experimental Molecular Medicine (CEMM), Amsterdam Infection and Immunity Institute, Meibergdreef 9, 1105 AZ, Amsterdam, the Netherlands; Amsterdam UMC, University of Amsterdam, Center for Experimental Molecular Medicine (CEMM),, Amsterdam UMC, Amsterdam Infection and Immunity Institute, Meibergdreef 9, 1105 AZ, Amsterdam, the Netherlands; Amsterdam UMC, University of Amsterdam, Department of Occupational Health and Safety, Meibergdreef 9, 1105 AZ, Amsterdam, the Netherlands; Amsterdam UMC, University of Amsterdam, Department of Medical Microbiology & Infection Prevention, Amsterdam Infection and Immunity Institute, Meibergdreef 9, 1105 AZ, Amsterdam, the Netherlands; Amsterdam UMC, University of Amsterdam, Department of Internal Medicine, Division of Infectious Diseases, Amsterdam Infection and Immunity Institute, Meibergdreef 9, 1105 AZ, Amsterdam, the Netherlands; Amsterdam UMC, Vrije Universiteit Amsterdam, Department of Internal Medicine, Division of Infectious Diseases, Amsterdam Infection and Immunity Institute, De Boelelaan 1117, 1081 HV, Amsterdam

## Abstract

**BACKGROUND:** It is unclear how, when and where health care workers (HCW) working in hospitals are infected with SARS-CoV-2.

**METHODS:** Prospective cohort study comprising 4-weekly measurement of SARS-CoV-2 specific antibodies and questionnaires from March to June 2020. We compared SARS-CoV-2 incidence between HCW working in Covid-19 patient care, HCW working in non-Covid-19 patient care and HCW not in patient care. Phylogenetic analyses of SARS-CoV-2 samples from patients and HCW were performed to identify potential transmission clusters.

**RESULTS:** We included 801 HCW: 439 in the Covid-19 patient care group, 164 in the non-Covid-19 patient care group and 198 in the no patient care group. SARS-CoV-2 incidence was highest in HCW working in Covid-19 patient care (13.2%), as compared with HCW in non-Covid-19 patient care (6.7%, hazard ratio [HR] 2.2, 95% confidence interval [CI] 1.2 to 4.3) and in HCW not working in patient care (3.6%, HR 3.9, 95% CI 1.8 to 8.6). Within the group of HCW caring for Covid-19 patients, SARS-CoV-2 cumulative incidence was highest in HCW working on Covid-19 wards (25.7%), as compared with HCW working on intensive care units (7.1%, HR 3.6, 95% CI 1.9 to 6.9), and HCW working in the emergency room (8.0%, HR 3.3, 95% CI 1.5 to 7.1). Phylogenetic analyses on Covid-19 wards identified multiple potential HCW-to-HCW transmission clusters while no patient-to-HCW transmission clusters were identified.

**CONCLUSIONS:** HCW working on Covid-19 wards are at increased risk for nosocomial SARS-CoV-2 infection, with an important role for HCW-to-HCW transmission.

(Funded by the Netherlands Organization for Health Research and Development ZonMw & the Corona Research Fund Amsterdam UMC; Netherlands Trial Register number NL8645)

## Introduction

In 2020 health care institutions worldwide were overwhelmed by Covid-19 patients. Stringent infection prevention and control measures have been applied to prevent transmission from patients to health care workers (HCW) and from HCW-to-HCW. Nonetheless, HCW have become infected during provision of care for COVID-19 patients and there is ongoing debate on which infection prevention and control measures are adequate.^1–3^ Delivering direct care to Covid-19 patients has been associated with infection or Covid-19 related hospital admission in some^4–8^ but not all studies. ^9–13^ Most studies were cross-sectional and retrospective, and lacked predefined control groups or detailed information on SARS-CoV-2 exposure including use of personal protective equipment (PPE).

To quantify the incidence of SARS-CoV-2 infection in HCW, identify potential risk factors and elucidate potential transmission routes, we performed the Serologic Surveillance of SARS-CoV-2 infection in health care workers (S3) study in two tertiary care medical centers in the Netherlands during the ‘first wave’ of SARS-CoV-2-infections. Serial serologic measurements were combined with phylogenetic analysis of viruses isolated from patients and HCW to identify transmission clusters.

## Methods

### Study design and population

We conducted a prospective serologic surveillance study in HCW of the Amsterdam University Medical Centers, the Netherlands, comprising two tertiary care hospitals. Four-weekly measurements of SARS-CoV-2 specific antibodies were performed over a period of 18 weeks during the first Covid-19 wave (March 23 - June 25, 2020). The first confirmed Covid-19 patient was admitted on March 9. Enrollment of HCW took place between March 23 and April 7, except for HCW in non-Covid-19 care who were enrolled during the final measurement in June 2020. Phlebotomies were combined with surveys including questions on personal and work-related SARS-CoV-2 exposure and symptoms. HCW were recruited by leaflets distributed in relevant departments with potentially eligible HCW and by intranet news items.

HCW were eligible for inclusion in one of three specific groups based on Covid-19 patient exposure: 1. HCW working as nurse or physician with bedside contacts with Covid-19 patients on a designated regular care Covid-19 ward, emergency room or intensive care unit; 2. HCW working as nurse or physician on a ward designated for non-Covid-19 care; and 3. HCW not in patient care. The second group participated only in the final measurement. The study was approved by institutional review boards of both hospitals, and written informed consent was obtained from each participant.

### Infection prevention practices

Both tertiary care centers instituted identical infection prevention and control measures in accordance with European and national guidelines.^14,15^ Initially, all HCW caring for (suspected) Covid-19 patients used PPE comprising disposable non-sterile gloves, gowns, FFP2 masks, and reusable goggles. From March 16 onwards, national guidelines on PPE were adjusted in accordance with recommendations at that time:^14,15^ HCW used type IIR surgical masks during non-aerosol generating care, and FFP2 masks on the intensive care and during high-risk, aerosol generating procedures. No PPE was recommended outside direct Covid-19 patient care, but social distancing measures were implemented hospital wide. Additional details regarding infection practices are provided in the Supplementary Appendix.

### Procedures

We collected survey data using Castor EDC.^16^ A survey example is provided in the online supplement. At each measurement, participants reported results of any preceding SARS-CoV-2 nucleic acid amplification test (NAAT) of nasopharyngeal swabs, performed as part of routine hospital screening of symptomatic HCW. SARS-CoV-2 specific antibodies were measured in serum using the Wantai SARS-CoV-2 pan-Ig anti-S1-RBD test according to manufacturer’s instructions (Beijing Wantai ELISA, Bioscience Co. (Chongqing) CLIA, Zuhai Livzon ELISA).^17^ Indeterminate results were classified as negative.

### Outcomes

Primary outcome was cumulative incidence of and time to SARS-CoV-2 infection during the study period. SARS-CoV-2 infection was defined as presence of SARS-CoV-2 specific antibodies above the threshold set by the manufacturer. Date of SARS-CoV-2 infection was defined as the sampling date of a first positive NAAT result or, in its absence, the midpoint in time between the last seronegative and the first seropositive sample. All participants were assumed to be seronegative on February 27 which was 4 weeks before the first measurement and the day the first Covid-19 patient was diagnosed in the Netherlands.

Outcomes were compared among the three study groups with varying levels of exposure to Covid-19 patients. Subgroup analysis included comparisons between hospital unit types (Covid-19 ward, intensive care unit and emergency room) and profession (nurse, physician). Secondary outcomes included infection rates in relation to self-reported exposure to Covid-19 patients, household Covid-19 contacts and presence of Covid-19 related symptoms; and results of the phylogenetic analyses.

### Statistical analysis

We used Kaplan-Meier estimates with log-rank test, and univariable and multivariable cox regression analyses to compare SARS-CoV-2 infection over time between study groups. The proportional hazard assumption did not hold because of fluctuating incidence of Covid-19 during the study period, evidenced by Schoenfeld tests resulting in p<0.05. The reported hazard ratios should therefore be interpreted as an average relative hazard for the entire study period, instead of a relative hazard at each individual time point. Multivariable models contained all other covariates used in the univariable models which were selected based on clinical relevance. Analysis was based on cases with complete data on covariates included in the regression models.

### Viral sequencing and phylogenetic analyses

To identify possible transmission clusters, virus sequencing was performed from routinely stored nasopharyngeal swabs of 26 infected HCW (not only including study participants) and 39 Covid-19 patients, selected from Covid-19 wards with high incidence of infection amongst HCW from which the biggest number of temporally related patient samples were also available. Included HCW worked on Covid-19 wards between March 15 and May 15; included patients had been admitted to corresponding wards between March 13 and April 19. Complete viral genomes were sequenced using the Ion AmpliSeq SARS-CoV-2 Panel, Ion Chef, and Ion Torrent S5 platforms (all Thermo-Fisher). Consensus full length SARS-CoV2 genomes (>29,000 nucleotide bases long with >100 minimum depth of coverage for each site) were generated by removing reads ends with PHRED scores <20 using Trimmomatic and mapping raw reads against the WIV04 reference genome (Genbank reference MN996528.1) using Bowtie 2.^18–20^

We used MAFFT (v7.427) to align SARS-CoV-2 sequences from HCW and patients, together with 300 randomly selected, contemporaneous SAR-CoV-2 virus genomes from the Netherlands (GISAID, Supplementary Data for accession numbers).^21^ We inferred a maximum likelihood tree with IQ-TREE (v2.0.6) using the HKY+I+G model.^22^ We applied Phydelity to the maximum likelihood tree to infer putative transmission clusters.^23^

We used BEAST (v1.10.4) to reconstruct a Bayesian time-scaled phylogenetic tree for the same set of sequences using the HKY+I+G model with a strict molecular clock, exponential growth prior, and an informative clock prior based on recent estimates of SARS-CoV-2 substitution rate (Γ-distribution prior with a mean of 0.8 × 10^**–3**^ subs/site/year and standard deviation of 5 × 10^**–4**^.^24,25^ We performed and combined two chains of 100 million steps. Convergence was reached for all parameters (ESS>700).

## Results

### Participants

We included 801 HCW: 439 in the Covid-19 patient care group, 164 in the non-Covid-19 patient care group and 198 in the no patient care group. Median age was 36 years (interquartile range [IQR] 29-50), 76% were female. HCW in Covid-19 and non-Covid-19 patient care were younger than HCW not working in patient care (median 34 years, IQR 29-44, and 33 years, IQR 27-49 versus 49 years, IQR 40-57, respectively). For measurements 2-4, survey completion rates were higher than the rate of HCW complying with blood sampling, likely because the former did not require physical presence.None of the participants with a SARS-CoV-2 infection reported being hospitalized during the study period. (Table 1).

**Table 1:** General characteristics of participants overall (A) and per measurement (B)

| **A: general characteristics of participants & survey results** |  |  |  |  |
| --- | --- | --- | --- | --- |
|  |  | **Covid-19 patient care (N=439)** | **non-Covid-19 patient care (N=164)** | **no patient care (N=198)** |
| age, median (interquartile range) |  | 37 (408-34) | 37 (164-33) | 47 (191-49) |
| sex | female | 289 (70) | 145 (88) | 146 (76) |
| position | nurse | 219 (50) | 129 (79) | 0 (0) |
|  | resident | 107 (24) | 25 (15) | 0 (0) |
|  | specialist | 86 (20) | 10 (6) | 0 (0) |
|  | other patient care | 27 (6) | 0 (0) | 0 (0) |
|  | administration/policy | 0 (0) | 0 (0) | 62 (31) |
|  | scientist | 0 (0) | 0 (0) | 43 (22) |
|  | pharmacy | 0 (0) | 0 (0) | 17 (9) |
|  | other non patient care | 0 (0) | 0 (0) | 76 (38) |
| tertiary care center | Amsterdam UMC, location AMC | 253 (58) | 73 (45) | 84 (42) |
|  | Amsterdam UMC, location VUmc | 186 (42) | 91 (55) | 114 (58) |
| days/week spent in hospital, mean (min-max) |  | 4.1 (1-5.5) | 3.8 (2-5.5) | 2.9 (1-5.5) |
| PCR test result | ever positive | 27 (6) | 7 (4) | 0 (0) |
|  | always negative | 165 (38) | 58 (35) | 20 (10) |
|  | never tested | 247 (56) | 99 (60) | 178 (90) |
| PPE training followed | E-learning only | 160 (37) | 65 (40) | - |
|  | simulation only | 13 (3) | 3 (2) | - |
|  | both | 253 (58) | 92 (56) | - |
|  | none | 11 (3) | 4 (2) | - |
| feasibility social distancing | easy | 2 (0) | 11 (7) | 42 (22) |
|  | medium | 27 (6) | 11 (7) | 56 (29) |
|  | difficult | 111 (27) | 46 (28) | 69 (36) |
|  | virtually impossible | 278 (67) | 96 (59) | 25 (13) |
| worried of getting Covid-19 | not at all | 117 (34) | 102 (62) | 52 (30) |
|  | somewhat | 164 (48) | 37 (23) | 92 (53) |
|  | medium | 53 (15) | 16 (10) | 26 (15) |
|  | very | 11 (3) | 9 (5) | 5 (3) |
| - if worried, most worried about | personal health | 53 (23) | 17 (27) | 28 (23) |
|  | infecting friends/family | 156 (69) | 40 (65) | 90 (73) |
|  | infecting patients | 18 (8) | 5 (8) | 0 (0) |
|  | infecting colleagues | 0 (0) | 0 (0) | 5 (4) |
| **B: character** |  |  |  |  |

| eristics<br>per<br>measur<br>ement |  |  |  |  |  |  |  |  |  |  |
| --- | --- | --- | --- | --- | --- | --- | --- | --- | --- | --- |
|  |  | **Covi**<br>**d-19**<br>**patie**<br>**nt**<br>**care** |  |  |  | **non-**<br>**Covi**<br>**d-19**<br>**patie**<br>**nt**<br>**care** | **no**<br>**patie**<br>**nt**<br>**care** |  |  |  |
|  | **Measure**<br>**ment #** | **1** | **2** | **3** | **4** | **4** | **1** | **2** | **3** | **4** |
| survey<br>complet<br>ed |  | 439<br>(100) | 411<br>(94) | 388<br>(88) | 349<br>(79) | 164<br>(100) | 198<br>(100) | 192<br>(96) | 184<br>(93) | 175<br>(88) |
| serology<br>availabl<br>e |  | 439<br>(100) | 367<br>(84) | 343<br>(78) | 288<br>(66) | 164<br>(100) | 197<br>(99) | 177<br>(89) | 172<br>(87) | 163<br>(82) |
| commun<br>ity<br>contact<br>(suspect<br>ed)<br>Covid-19<br>patients | household | 18 (4) | 16 (4) | 8 (2) | 1 (0) | 2 (1) | 3 (2) | 3 (2) | 0 (0) | 1 (1) |
|  | other | 24 (5) | 12 (3) | 6 (2) | 6 (2) | 11 (7) | 4 (2) | 4 (2) | 2 (1) | 2 (1) |
|  | none | 397<br>(90) | 380<br>(93) | 373<br>(96) | 338<br>(98) | 151<br>(92) | 191<br>(96) | 184<br>(96) | 182<br>(99) | 172<br>(98) |
| severity<br>of<br>Covid-19-like<br>syndrom<br>e | no<br>symptoms | 249<br>(61) | 306<br>(75) | 319<br>(82) | 291<br>(84) | 98<br>(60) | 147<br>(77) | 160<br>(84) | 161<br>(88) | 152<br>(87) |
|  | minimal<br>(i.e. no<br>limitation<br>s in daily<br>functionin<br>g) | 112<br>(27) | 81<br>(20) | 57<br>(15) | 41<br>(12) | 46<br>(28) | 28<br>(15) | 27<br>(14) | 17 (9) | 20<br>(11) |
|  | mild (i.e.<br>some<br>limitation<br>s in daily<br>functionin<br>g) | 14 (3) | 12 (3) | 4 (1) | 10 (3) | 6 (4) | 6 (3) | 4 (2) | 5 (3) | 2 (1) |
|  | moderate<br>(i.e. most<br>of the day<br>supine) | 33 (8) | 9 (2) | 7 (2) | 4 (1) | 14 (9) | 9 (5) | 0 (0) | 1 (1) | 1 (1) |
| hospital<br>unit<br>type | intensive<br>care unit | 148<br>(34) | 170<br>(41) | 118<br>(30) | 49<br>(14) | 0 (0) | - | - | - | - |
|  | Covid-19 unit | 106 (24) | 120 (29) | 69 (18) | 41 (12) | 0 (0) | - | - | - | - |
|  | emergency room | 97 (22) | 93 (23) | 93 (24) | 87 (25) | 0 (0) | - | - | - | - |
|  | combination of above | 88 (20) | 0 (0) | 0 (0) | 0 (0) | 0 (0) | - | - | - | - |
|  | non-Covid-19 ward | 0 (0) | 28 (7) | 108 (28) | 173 (49) | 164 (100) | - | - | - | - |
| PPE always correctly used |  | 405 (94) | 363 (96) | 266 (96) | 153 (96) | 162 (99) | - | - | - | - |
| # visits (suspected) Covid-19 patient room | >50 times | 107 (24) | 138 (34) | 44 (11) | 8 (2) | 0 (0) | - | - | - | - |
|  | 26-50 times | 40 (9) | 64 (16) | 36 (9) | 12 (3) | 0 (0) | - | - | - | - |
|  | 11-25 times | 109 (25) | 83 (20) | 63 (16) | 20 (6) | 0 (0) | - | - | - | - |
|  | 6-10 times | 54 (12) | 59 (14) | 54 (14) | 41 (12) | 0 (0) | - | - | - | - |
|  | 1-5 times | 89 (20) | 37 (9) | 79 (20) | 78 (22) | 164 (100) | - | - | - | - |
|  | none | 38 (9) | 29 (7) | 111 (29) | 188 (54) | 0 (0) | - | - | - | - |

*Caption:* PPE, personal protective equipment

### Primary outcome

SARS-CoV-2 cumulative incidence was highest in HCW working in Covid-19 patient care (13.2%), as compared with HCW in non-Covid-19 patient care (6.7%, hazard ratio [HR] 2.2, 95% confidence interval [CI] 1.2 to 4.3) and in HCW not working in patient care (3.6%, HR 3.9, 95% CI 1.8 to 8.6, Figure 1A). Within the group of HCW caring for Covid-19 patients, SARS-CoV-2 cumulative incidence was highest in HCW working on Covid-19 wards (25.7%), as compared with HCW working on intensive care units (7.1%, HR 3.6, 95% CI 1.9 to 6.9), and HCW working in the emergency room (8.0%, HR 3.3, 95% CI 1.5 to 7.1, Figure 1B. Figure 1C shows the number of Covid-19 admissions to study hospitals and regional Covid-19 incidence. Results were similar for individual study sites (Figures S1A+B & S2A+B in the Supplementary Appendix) and when including either only NAAT or only serology results in the analysis (Figures S1C+D & S2C+D in the Supplementary Appendix).Main results were similar after adjustment in the multivariable cox regression. Contact with a Covid-19 positive person in the community (including household) and contact with a Covid-19 positive coworker were associated with Covid-19 infection (Table 2).

**Table 2:** Results of univariable and multivariable cox regression analysis of association between SARS-CoV-2 infection and determinants in the overall study population (A) and within Covid-19 patient care (B)

| **A: univariable and multivariable cox regression models** |  |  |  |  |
| --- | --- | --- | --- | --- |
|  |  | SARS-CoV-2 incidence<br>N/total (% , 95% CI)) | HR (95%<br>CI) | Adjusted<br>HR (95%<br>CI) |
| HCW work<br>environment | no patient care | 7/198 (3.6, 0.9-6.1) | 1 | 1 |
|  | non-COVID-19<br>patient care only | 11/164 (6.7, 2.8-10.5) | 1.7 (0.7-4.5) | 1.6 (0.6-4.4) |
|  | COVID-19 patient<br>care | 54/439 (13.2, 9.9-16.4) | 3.9 (1.8-8.6)* | 3.1 (1.2-7.7)* |
| COVID-19<br>coworker contact | no | 30/455 (7.0, 4.5-9.4) | 1 | 1 |
|  | yes | 40/319 (13.5, 9.5-17.3) | 2.0 (1.3-3.2) | 1.7 (0.99-3.0) |
| COVID-19<br>community contact | no | 52/693 (8.2, 6.0-10.3) | 1 | 1 |
|  | yes | 20/108 (20.2, 11.8-27.9) | 2.6 (1.6-4.3) | 2.0 (1.1-3.6) |
| days/week spent in<br>hospital | - | - | 1.1 (0.8-1.3) | 1.7 (0.99-3.0) |
| age | - | - | 0.98<br>(0.96-0.997) | 0.99 (0.97-1.01) |
| **B: univariable and multivariable cox regression models within COVID-19 patient care group** |  |  |  |  |
|  |  | SARS-CoV-2 incidence<br>N/total (% , 95% CI) | HR (95%<br>CI) | Adjusted<br>HR (95%<br>CI) |
| hospital unit type | intensive care | 13/186 (7.1, 3.3-10.7) | 1 | 1 |
|  | COVID-19 unit | 32/134 (25.7, 17.6-33.1) | 3.6 (1.9-6.9)** | 3.7 (1.7-8.3)** |
|  | emergency room | 7/102 (8.0, 2.5-13.1) | 1.1 (0.5-2.7) | 1.3 (0.5-3.6) |
|  | combination of above | 2/17 (11.8, 0.0-25.8) | 2.0 (0.5-9.0) | 1.2 (0.1-11.8) |
| position | specialist | 5/86 (6.4, 0.7-11.8) | 1 | 1 |
|  | resident | 14/107 (14.7, 7.5-21.3) | 2.7 (0.98-7.4) | 1.7 (0.5-5.7) |
|  | nurse | 35/246 (14.9, 10.2-19.3) | 2.6 (1.01-6.6) | 1.6 (0.6-4.6) |
| self-reported COVID-19 exposure | low | 13/88 (15.3, 7.3-22.7) | 1 | 1 |
|  | medium | 16/165 (11.2, 6.0-16.1) | 0.7 (0.3-1.4) | 0.5 (0.2-1.2) |
|  | high | 13/73 (18.2, 8.7-26.6) | 1.1 (0.5-2.4) | 1.1 (0.4-2.9) |
|  | very high | 12/113 (10.8, 4.8-16.4) | 0.7 (0.3-1.4) | 0.8 (0.3-2.1) |
| feasibility social distancing | easy | 0/2 (0.0, 0.0-0.0) | NA | NA |
|  | medium | 7/27 (26.7, 7.5-41.8) | 1 | 1 |
|  | difficult | 19/111 (18.5, 10.5-25.8) | 0.6 (0.3-1.4) | 0.4 (0.1-1.3) |
|  | virtually impossible | 26/278 (9.9, 6.3-13.4) | 0.3 (0.1-0.7) | 0.3 (0.1-0.9) |
| PPE always correctly used | no | 5/54 (9.4, 1.2-16.9) | 1 | 1 |
|  | yes | 49/383 (13.8, 10.2-17.3) | 1.4 (0.6-3.6) | 1.2 (0.4-3.5) |
| COVID-19 coworker contact | no | 17/186 (9.3, 5.0-13.5) | 1 | 1 |
|  | yes | 35/232 (16.0, 11.1-20.7) | 1.7 (0.96-3.0) | 2.4 (1.1-5.0) |
| COVID-19 community contact | no | 39/360 (11.8, 8.2-15.1) | 1 | 1 |
|  | yes | 15/79 (19.7, 10.2-28.2) | 1.8<br>(0.996-3.3) | 1.5 (0.7-3.1) |
| days/week spent in hospital | - | - | 0.7 (0.5-0.9) | 0.7 (0.5-0.9) |
| age, years | - | - | 0.97<br>(0.95-1.002) | 1.00 (0.97-1.03) |

*Caption:* HR, hazard ratio; CI, confidence interval. Percentages with confidence intervals were calculated using the Kaplan Meier method. Adjusted HRs shown in A belong to a model containing all variables in A, likewise for B. \*when compared to non-COVID-19 patient care only: HR 2.2, 95% CI 1.2-4.3, adjusted HR 1.9, 0.98-3.8, \*\* when compared to emergency room: HR 3.3, 95% CI 1.5-7.1, adjusted HR 2.9, 95% CI 1.2-7.1.

**Figure 1.**
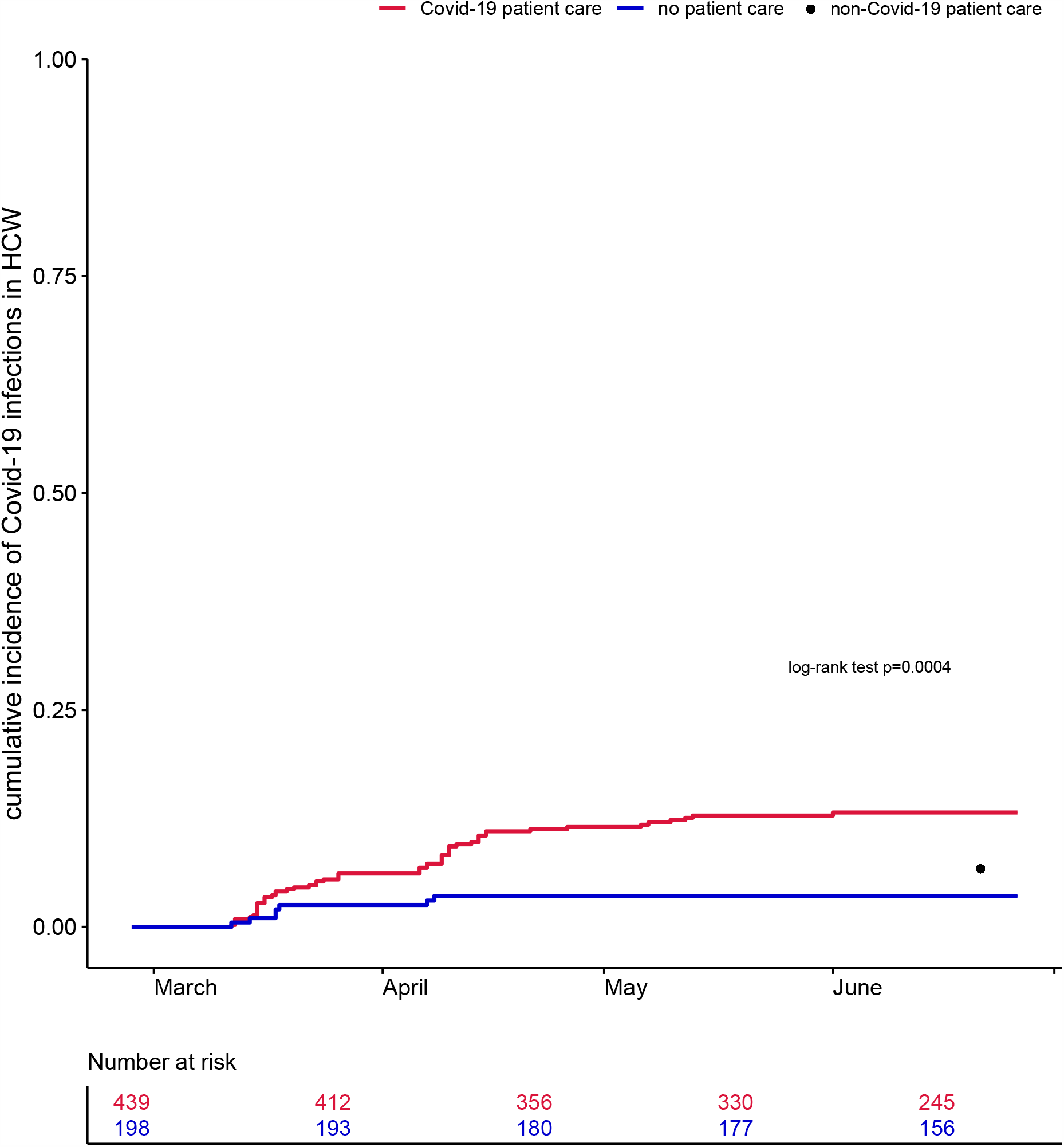
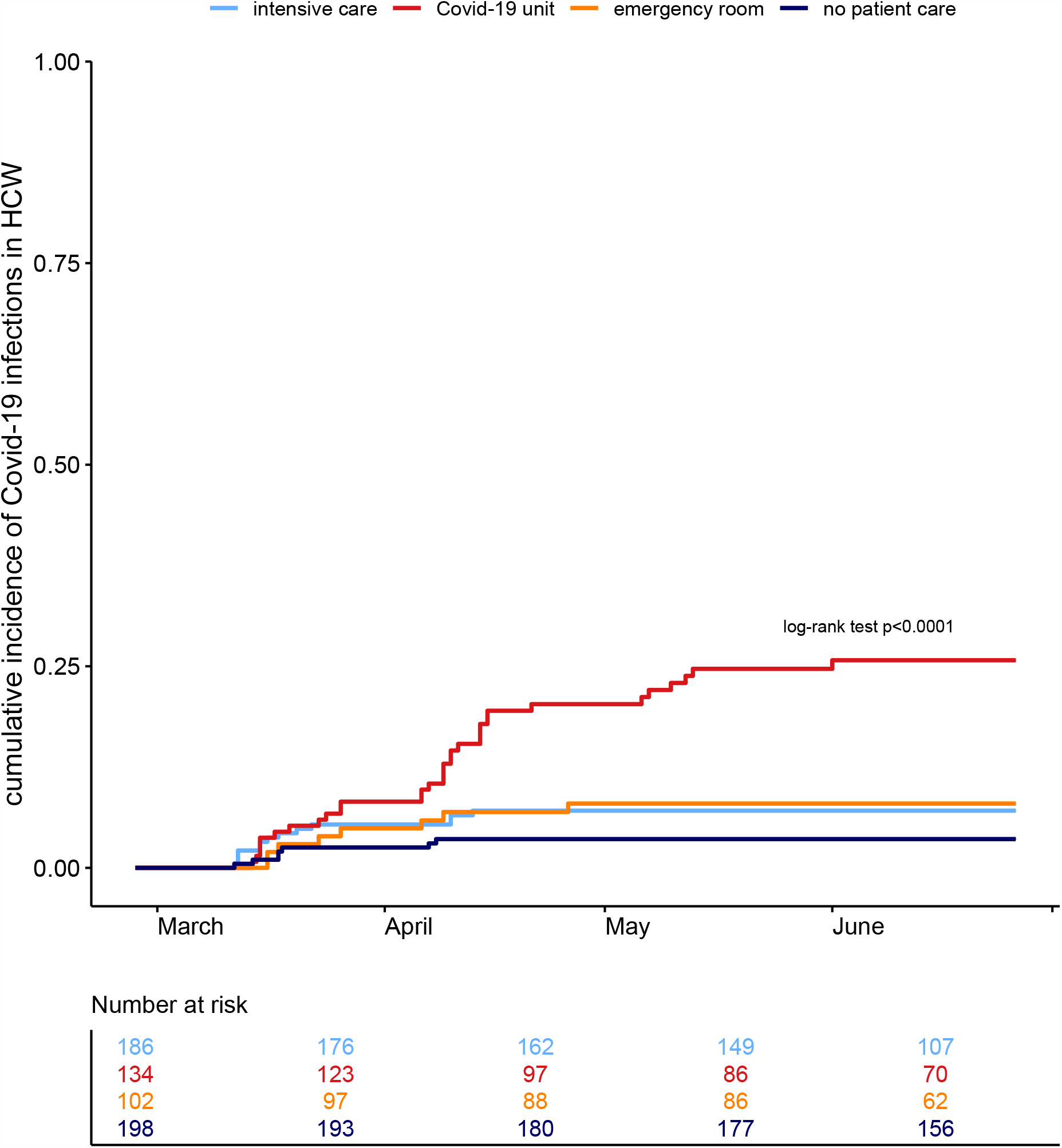
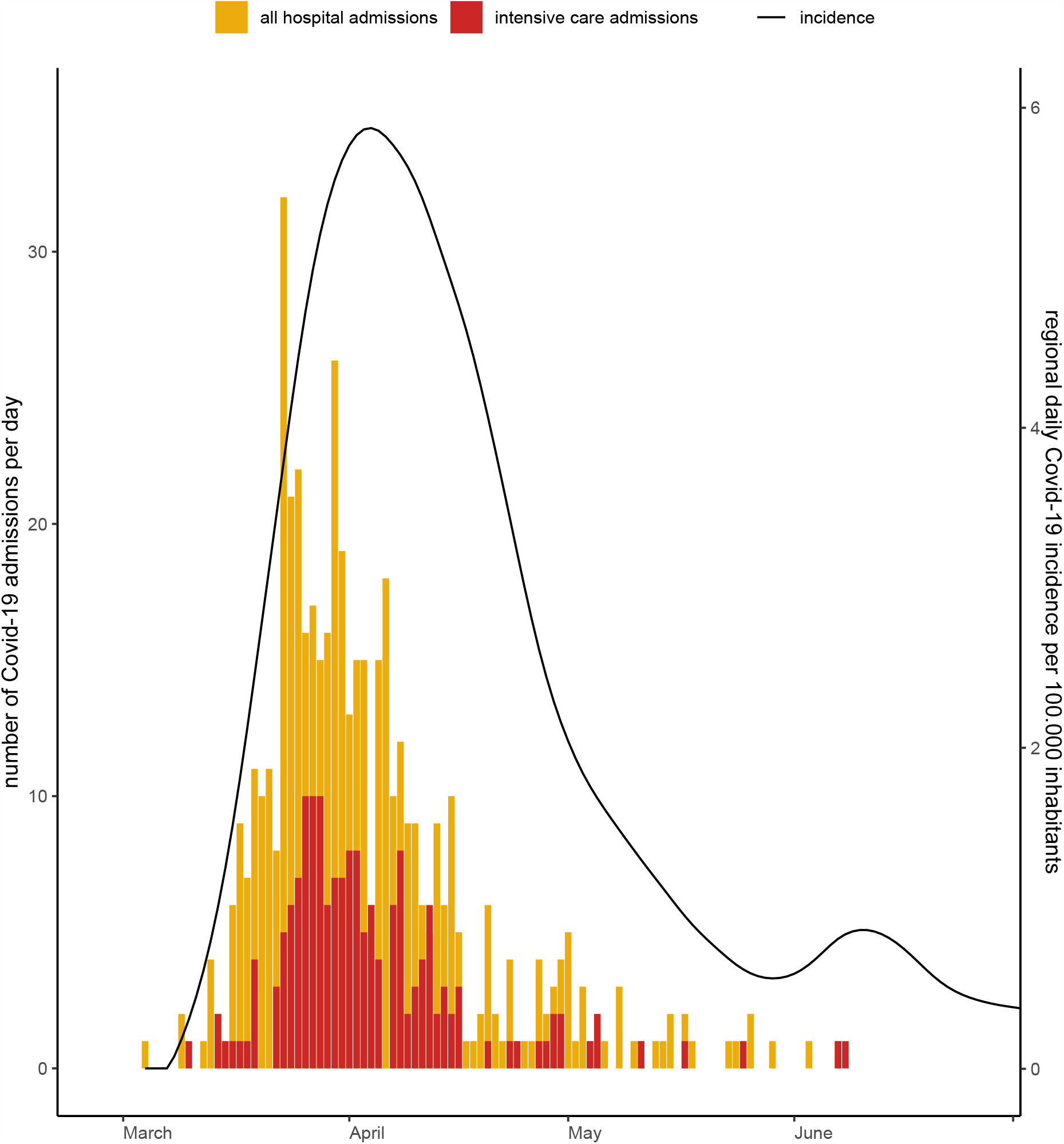
A: Cumulative incidence of SARS-CoV-2 infection in health care workers with different levels of Covid-19 patient exposure B: Cumulative incidence of SARS-CoV-2 infection in health care workers defined by hospital unit type C: Number of Covid-19 hospital and intensive care admissions, and regional Covid-19 incidence A+B: Date of SARS-CoV-2 infection was defined as the sampling date of a first positive nucleic acid amplification test result or, in its absence, the midpoint in time between the last seronegative and the first seropositive sample. All participants were assumed to be seronegative on February 27 which was 4 weeks before the first measurement and the day the first Covid-19 patient was diagnosed in the Netherlands. A: Log-rank test reflects differences between health care workers (HCW) in Covid-19 care and HCW not in patient care (no patient care). HCW in non-Covid-19 patient care (black dot) were included in the fourth measurement only and added to the figure for reference. B: Log-rank test reflects differences between all HCW groups shown. Participants working on multiple hospital unit types during the study were excluded from this analysis because of small group size (N=17). ICU, intensive care unit. C: Left Y-axis: number of patients with Covid-19 admitted to both hospital sites (yellow bars) or intensive care units (red bars). Source: hospital administrative records, assembled with help from the CovidPredict research initiative: https://covidpredict.org. Right Y-axis: incidence of Covid-19 in the Dutch province Noord Holland where both hospital sites are located. Source: National Institute for Public Health and the Environment, the Netherlands.

Among HCW working in Covid-19 care, cumulative incidence among physicians was 11.0%; specialists had lower cumulative incidence than residents (6.4% as compared with 14.7%, HR 2.7 95% CI 0.98 to 7.4) and nurses (14.9%, HR 2.6 95% CI 1.01 to 6.6).

SARS-CoV-2 incidence in HCW was particularly high on one regular Covid-19-ward compared to other Covid-19 wards (ward 2; Figure S3 in the Supplementary Appendix). This ward was similar to the other wards with regard to HCW deployment and architectural structure, but had a higher proportion of patients with pre-existing pulmonary disease and use of high-flow nasal oxygen therapy. To assess contribution of this ward to overall results, the primary outcome was reanalyzed when excluding this ward, resulting in a SARS-CoV-2 incidence of HCW on Covid-19 units of 19.7% (compared with HCW on the intensive care units: HR 2.8, 95% CI 1.4 to 5.5, Table S1 in the Supplementary Appendix).

### Secondary outcomes

Of the 72 participants with seroconversion, 33 participants (45%) also tested positive by NAAT during routine screening of symptomatic HCW, all of which were HCW in direct patient care due to the restrictive access to SARS-CoV-2 testing at that time. Only one participant without documented seroconversion tested positive by NAAT, which occurred prior to the fourth measurement, but the subsequent blood sample was mislabeled and therefore not analyzed.

HCW with SARS-CoV-2 infection reported at least one symptom suggestive of Covid-19 (cough, headache, sore throat, fever, dyspnea, chest pain, anosmia, cold, diarrhea) in 85% of cases, compared to 86% of participants without infection. After adjustment for all other symptoms, only anosmia was associated with infection: 33/72 (54%) in seropositive participants compared to 14/729 (2%) in negative participants, adjusted HR 25.0, 95% CI 13.7 to 45.4.

### Phylogenetic analyses

In the maximum likelihood phylogeny, 32 out of 39 sequences from patients admitted to a Covid-19 ward (Figure 2A, triangle-shaped tips) and 12 of the 26 from HCW were dispersed across the tree among the 300 contemporaneous viruses from the Netherlands suggesting unrelated infections. Phydelity identified 5 putative transmission clusters containing the remaining 21 sequences (7 patients, 14 HCW, Figure 3A). Clusters A and B comprised patients clustering with each other or with HCWs. The three other transmission clusters (C, D, and E) contained only HCW.

**Figure 2:**
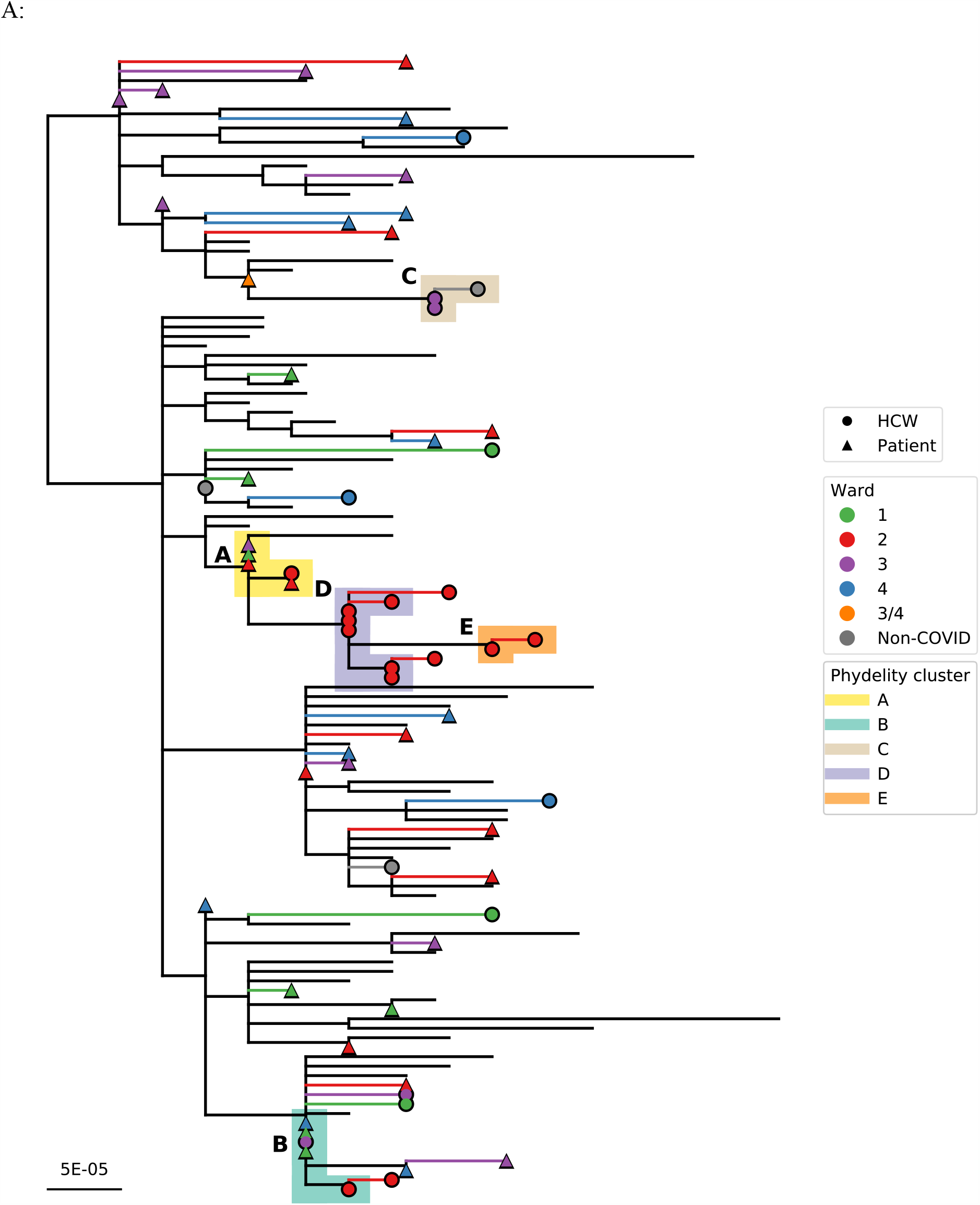
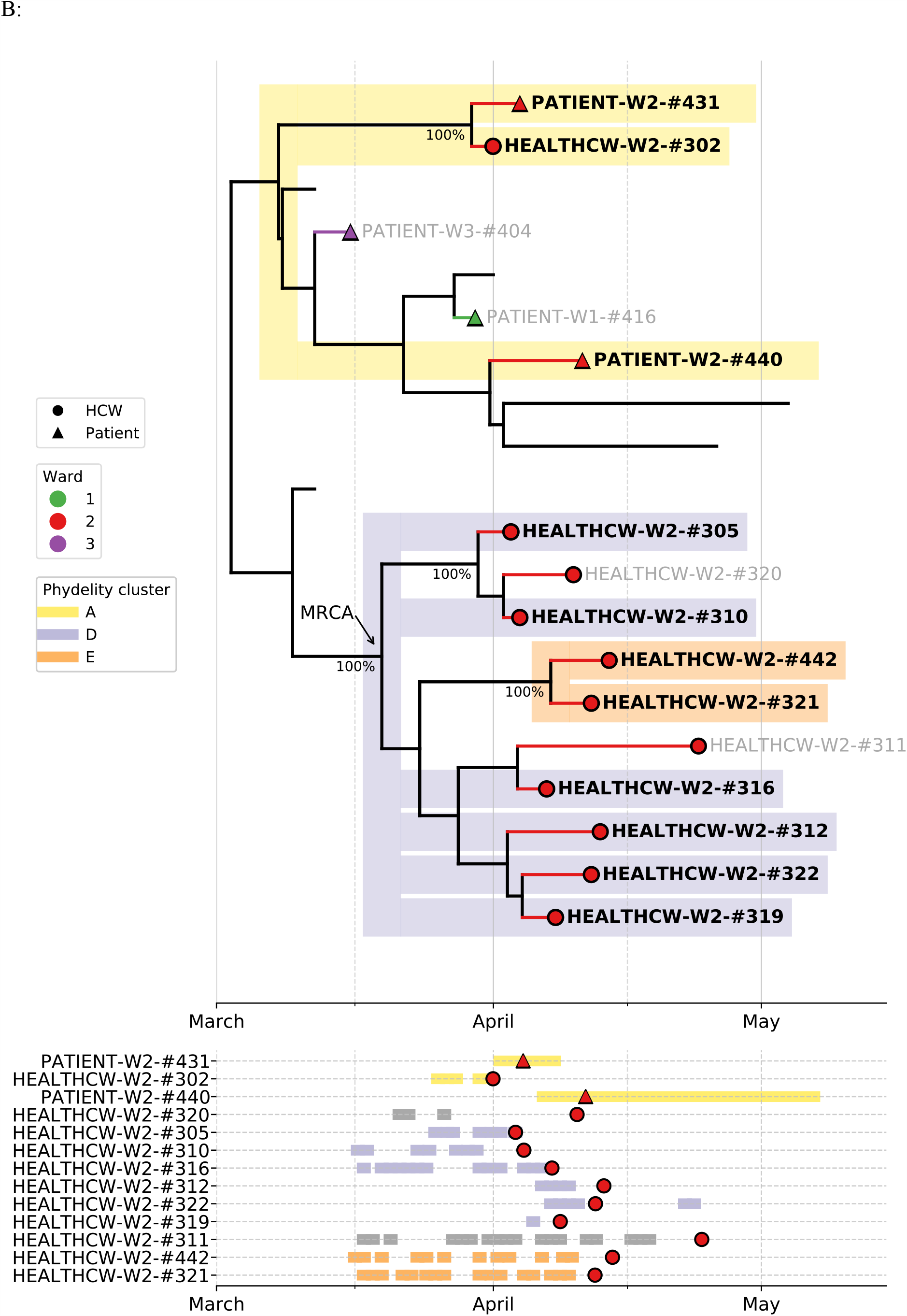
Maximum likelihood phylogeny of SARS-CoV-2 sequences with identified potential transmission clusters (A) and time-scaled subtree for clusters A, D, and E (B) A: A condensed maximum-likelihood phylogeny of SARS-CoV-2 sequences that were collected (marked with tip shapes) and a random sample of contemporaneous reference sequences (no tips) circulating within the Netherlands. Tip shapes are colored according to the wards the patients (triangle tips) and healthcare workers (HCW; circle tips) were assigned to. Potential transmission clusters identified by Phydelity are shaded by different colors. B: Top panel: Subtree of a time-scaled BEAST phylogeny that subtends sequences collected from individuals linked in potential transmission clusters A, D and E. Transmission clusters were identified based on the maximum likelihood tree (i.e. genetic similarity). Sequences that were collected in the study hospitals were marked with tip shapes, those not ending in a tip shape are contemporaneous reference sequences circulating within The Netherlands within the same time period. Tip shapes are colored according to the wards the patients (triangle tips) and HCW (circle tips) were assigned to. Internal nodes with >90% posterior support are annotated next to the node. The most recent common ancestor (MRCA) of individuals linked by transmission cluster D and E is also labelled. Bottom panel: Timeline of working shifts and admittance of HCWs and patients respectively. Red circle denotes the date of positive nucleic acid amplification test. This time-scaled tree gives more information if these clusters could have arisen from a single introduction (i.e. have a MRCA), and if so gives an estimate of when that might be.

Patient-to-patient and HCW-to-patient transmission is unlikely because patients admitted to the Covid-19 wards had NAAT proven or highly suspected SARS-CoV-2 infection based on symptoms or radiological findings at time of admission. This is further evidenced by the lack of clear epidemiological links between patients in clusters A and B. There was also no evidence of patient-to-HCW transmission based on our phylogenetic analysis and there was no overlap between the patient admission dates and HCW working shifts in clusters A and B (Supplementary Figure S4).

In the three clusters containing only HCWs, there was a high degree of overlap in working shifts suggesting epidemiological linkage. Two out of three clusters (D and E) contained only sequences obtained from HCWs working in ward 2. The time-scaled phylogeny (Figure 2B) suggests a single introduction for these HCWs working in ward 2 around mid-March (median date: March 19, 2020, 95% highest posterior density interval: March 11 – March 30, 2020; 100% posterior support).

## Discussion

We have prospectively followed a large cohort of HCW during the first wave of the Covid-19 pandemic with the aim of comparing cumulative SARS-CoV-2 incidence between groups of HCW with varying exposure to Covid-19 patients. Our results show a consistently higher risk of SARS-CoV-2 infection for HCW caring for Covid-19 patients, compared to HCW in non-Covid-19 patient care or HCW not working in patient care. Subgroup analysis shows the overall risk was largely driven by a substantially increased risk in HCW on regular care Covid-19 wards; infection rates in HCW working in the intensive care units and emergency room were comparable to HCW working in non-Covid-19 care. Our phylogenetic analysis combined with epidemiologic data identified transmission clusters comprising only HCW, consistent with HCW-to-HCW transmission on Covid-19 wards, while no evidence of patient-to-HCW transmission was found.

SARS-CoV-2 seroprevalence in HCW not working in patient care was comparable to healthy blood donors in the Dutch general population at the time.^26^ The higher incidence in HCW working in patient care of any kind suggests that working in patient care increases infection risk. Incidence of infection in HCW in Covid-19 care was highest, which could suggest that patient-to-HCW transmission was responsible for the excess incidence in this group. However, we did not find an association between infection and self-reported number of contacts with Covid-19, which would have been expected if patient-to-HCW transmission was the dominant transmission pattern. Additionally, on one Covid-19 ward (of six) multiple HCW were infected before the first Covid-19 patient was admitted. Finally, the phylogenetic analyses showed no evidence for patient-to-HCW transmission, although this cannot be completely ruled out.

Phylogenetic analyses showed evidence for HCW-to-HCW transmission on Covid-19 units. The hypothesis that HCW-to-HCW transmission plays an important role is further supported by the increased incidence among HCW who reported contact with a SARS-CoV-2 positive colleague. More than half of seropositive HCW in our study did not report a positive NAAT result, suggesting a significant proportion of infections in HCW remained unrecognized. As a result, HCW likely have been working whilst unaware of their SARS-CoV-2 infection, hence presenting a risk of transmission. The number of HCW present on Covid-19 wards was higher than on other regular care wards due to the nature of care and because mobility of HCW working in Covid-19 care through the hospital was discouraged. Personnel break rooms were therefore more crowded than usual. While universal masking was not yet recommended during this period, it is arguable whether this would have made a difference since masks cannot be worn while eating or drinking. The intensive care units differed with regard to facilitating social distancing by using additional break rooms with clearly demarcated spaces between seats.

Our study has a number of limitations. First, despite the prospective cohort design, selection bias cannot be completely ruled out, e.g. HCW staying at home ill were not able to enroll if this happened during the first measurement resulting in underestimating of incidence. Second, not all nasopharyngeal samples from patients and HCW collected for SARS-CoV-2 NAAT were available for viral sequencing analyses as they were either not stored or the admitted patients were diagnosed elsewhere. As such, there could be missing clusters and/or missing links in the transmission clusters that were inferred. Third, no systematic data on compliance to infection prevention measures were collected, limiting more precise conclusions. Fourth, infection incidence was substantially higher on one specific Covid-19 ward, which also contributed the majority of transmission clusters. However, when excluding this ward, the proportion of seroconverted HCW on regular Covid-19 wards remained more than double as high when compared to intensive care, emergency room or non-covid-19 wards.

Preventing SARS-CoV-2 infection in HCW is important for the health of the individual HCW, to halt the ongoing pandemic and to maintain a functioning healthcare system. Understandably, much attention has been focused on preventing patient-to-HCW transmission. Our results show that working in hospital patient care leaves HCW vulnerable to infection through HCW-to-HCW transmission, which has received less attention and deserves more consideration. We recommend in the current situation of high SARS-CoV2 incidence optimal measures to facilitate social distancing on the work floor, e.g. reducing the number of people per room by spreading break times, increasing size or number of break rooms, enabling online conferencing, universal use of face masks, and investing in structural auditing and training by infection prevention and control personnel.

In conclusion, HCW working on Covid-19 wards are at increased risk for nosocomial SARS-CoV-2 infection, with an important role for HCW-to-HCW transmission.

## Supporting information

GISAID acknowledgements

## Data Availability

Data are not made public at the moment. Please contact corresponding author for data requests.

## Acknowledgements

We thank all participating healthcare workers of Amsterdam UMC, who took time to facilitate this study in the midst of the pandemic, for their contributions. We thank Adinda Pijpers, Esmee Das, Nikita Borstlap and Lisa Urlings for their essential help in performing the study measurements.

