## Supplementary material for "Serologic Surveillance and Phylogenetic Analysis of SARS-CoV-2 Infection in Hospital Health Care Workers": Online supplement

### Supplemental methods

#### Infection prevention practices

At the peak of the epidemic, both hospitals accommodated three regular care Covid-19 wards (from now on referred as Covid-19 wards) in addition to Covid-19 intensive care units in both hospitals. Suspected and confirmed Covid-19 patients were cared for on dedicated Covid-19 wards in droplet and contact isolation, in addition to standard precautions. No visitors were allowed on Covid-19 wards except for those visiting terminally ill patients. From March 23 onwards Covid-19 patients could receive care in cohorts with a maximum of four patients per room. A short E-learning on donning and doffing PPE was mandatory for all HCW caring for patients. Attending a real-life practice simulation training was optional. All ventilation and air pressure conditions on Covid-19 wards were maintained according to national guidelines.(1) Infection control practitioners visited Covid-19 wards each working day during the study period to advice and answer questions of HCW. Until June 1st, SARS-CoV-2 NAAT was available only for symptomatic HCW involved in patient care in addition to suspected Covid-19 cases requiring hospital admission. After this date testing was available for all employees and patients with symptoms. suggestive of SARS-CoV-2 infection. There were no critical shortages of PPE in participating hospitals during the study period.

#### Statistical analysis

We set the start of survival time on February 27 (exactly 4 weeks before the start of this study), since on that day the first Covid-19 patient was diagnosed in the Netherlands.(32) Participants without SARS-CoV-2 infection were censored at the last day of the measurement in which they participated. Cox regression analysis was not performed, since the proportional hazards assumption was not met due to the uneven hazard during the study period. Inclusion targeted a sample size per group of at least 157 participants (including subgroups based on hospital unit type and profession), assuming alpha of 5%, to reach 80% power to detect a infection rate difference of 10% or more.(2) All analyses were performed using R version 4.0, and packages survminer and survival.(3–5)

**Example of survey (translated from Dutch)**

**1) Have you worked in patient care with COVID patients since the second measurement?**

-No

-Yes

If answer question 1 = No.

**2) What position have you held in the past 4 weeks?**

-Administrative assistant

-Policy officer

-Scientist/researcher

-Doctor/nurse outside of patient care

-Doctor / nurse on a ward without COVID patients

-ICT

-Laboratories

-Security

-Other

If answer question 2 = Other

**3) What position was that?**

……….

**4) How many days have you worked in the hospital in the past 4 weeks?**

……….

If answer question 1 = Yes

**5) In which hospital departments have you had direct contact with COVID patients since the second measurement? Multiple answers possible**.

-the emergency room

-the intensive care unit, location where corona patients are also treated

- Ward 1
- Ward 2
- Ward 3
- Ward 4

- Ward 5
- Ward 6
- other corona ward

If answer question 5 = other corona department

**6) Which department was this?**

……….

If answer question 1 = Yes.

**7) In which department have you had the most contact with COVID patients since the second measurement? Only 1 answer is possible here.**

-the emergency room

-the intensive care unit, location where corona patients are also treated

- Ward 1
- Ward 2
- Ward 3
- Ward 4
- Ward 5
- Ward 6
- other corona ward

**8) Which department was this?**

……….

If answer question 1 = Yes.

**9) How many times have you been in the room of a patient suspected or proven to be infected with the corona virus (SARS-CoV-2) since the second measurement estimated?**

-Not

-1-5 times

-6-10 times

-11-25 times

-26-50 times

-More than 50 times

If answer question 9 ≠ Not

**10) During your own contacts with the patients as mentioned in the previous question, have you always adhered to the infection prevention guidelines (isolation, protective clothing, etc.) as prescribed?**

-No

-Yes

**11) Have you had any symptoms since the second measurement? Multiple answers possible.**

-Fever (>38.5 degrees Celsius)

-Coug (>24 hours)

-Nasal cold (>24 hours)

-Diarrhea (3x a day or more) or vomiting

-Headache (>24 hours)

-Reduced smell and/or taste ( 24 hours)

-Sore throat (>24 hours)

-Dyspnea

-Chest pain

-None of the above

If answer question 11 ≠ None of the above

**12) What was approximately the first day of symptoms since the second measurement?**

DD-MM-YYYY

If answer question 11 ≠ None of the above

**13) The next question concerns the severity of symptoms on the day you felt the most ill since the second measurement. Choose the description that best suited your situation that day.**

-Despite the complaints I was able to fully perform all my normal activities (for example work and housekeeping).

-Due to the complaints I could no longer perform some normal activities (for example work and housekeeping).

-Due to the complaints, I spent most of the day in bed or on the couch.

-I was admitted to hospital because of the complaints.

If answer question 13 = I was admitted to hospital because of the complaints.

**14) In which department have you been admitted?**

-In a normal nursing ward

-In a corona ward

-In intensive care

**15) Have you been tested for infection with the corona virus (SARS-CoV-2) (by PCR throat / nose swab) since the second measurement?**

-No

-Yes

If answer question 15 = Yes

**16) How often have you been tested?**

……….

If answer question 15 = Yes

**17) Was (one of) the test (s) positive?**

-No

-Yes

If answer question 17 = Yes

**18) What was the approximate date of the first positive test?**

DD-MM-YYYY

If answer question 17 = No.

**19) What were the approximate dates on which the tests were negative? In case of multiple negative tests, please mention all dates here.**

……….

**20) Since the second measurement, have you worked with a colleague who had (mild) complaints at that time and who eventually tested positive for infection with the corona virus (SARS-CoV-2) (by means of PCR throat nose swab)?**

-No
-Yes

**21) Have you had contact outside of work (physical contact, or spent>1 hour in the same room) with someone who at that time had (mild) complaints and who eventually tested positive for infection with the corona virus (SARS-CoV- 2) (by PCR throat-nose wat) since the second measurement?**
-No
-Yes, it was a roommate
-Yes, it was a different person

If answer question 21 = Yes, it was a different person

**22) In what relationship were you with that person?**
……….
**23) Have you had contact outside of work (physical contact, or spent 1 hour in the same room) with someone who at that time had (mild) complaints with high suspicion of being infected with the coronavirus (SARS-CoV-2) since the second measurement**
-No
-Yes, it was a roommate
-Yes, it was a different person
If answer question 23 = Yes, it was a different person

**24) In what relationship were you with that person?**
……….

**25) How easy is it to stay 1.5 meters away from your direct colleagues at work?**

-Very easy
-Simple
-Neutral
-Difficult
-Virtually impossible

### Supplemental figures

#### Figure S1

A: Cumulative incidence of SARS-CoV-2 infection in health care workers with different levels of Covid-19 patient exposure for hospital 1

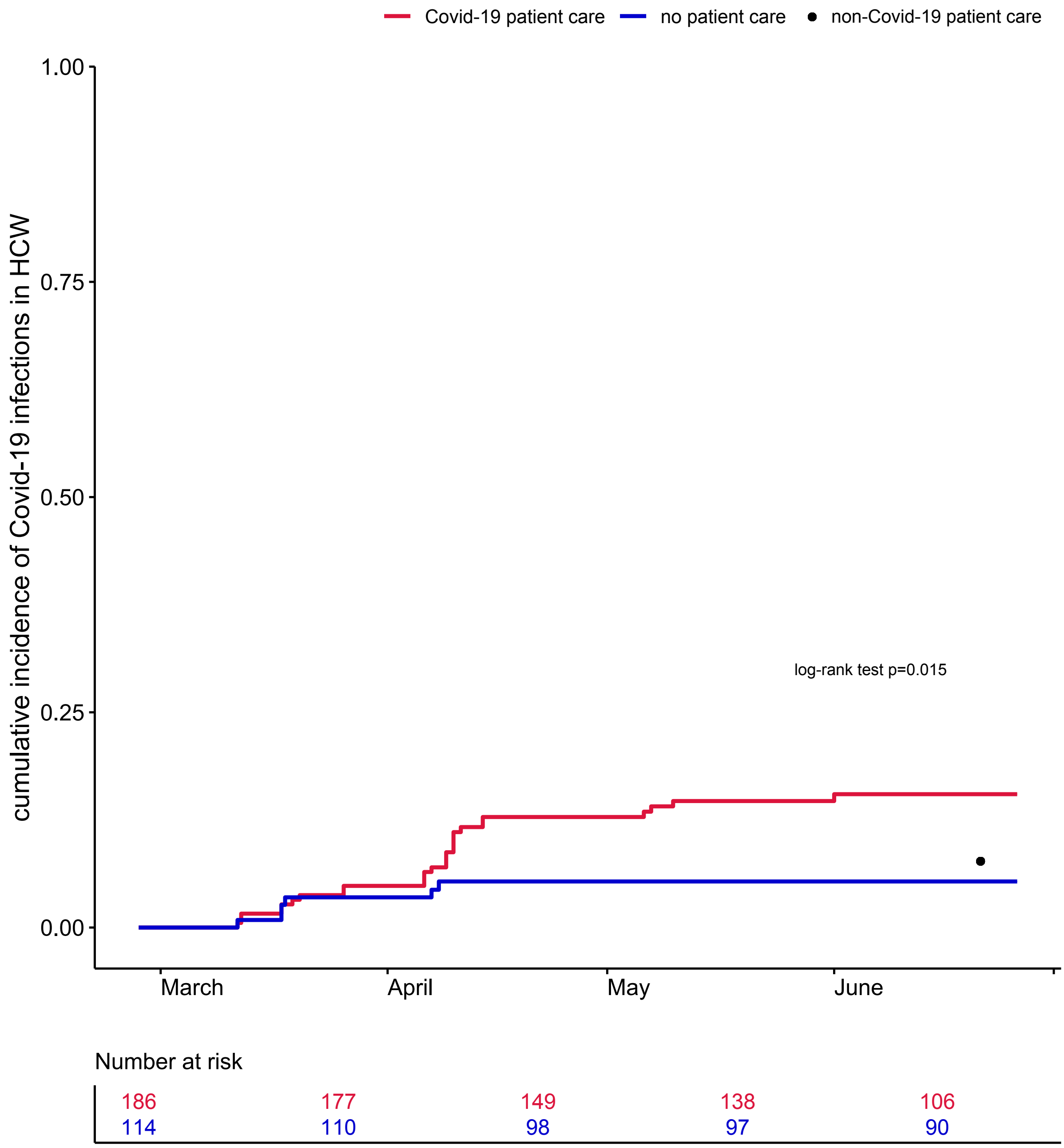

B: Cumulative incidence of SARS-CoV-2 infection in health care workers with different levels of Covid-19 patient exposure for hospital 2

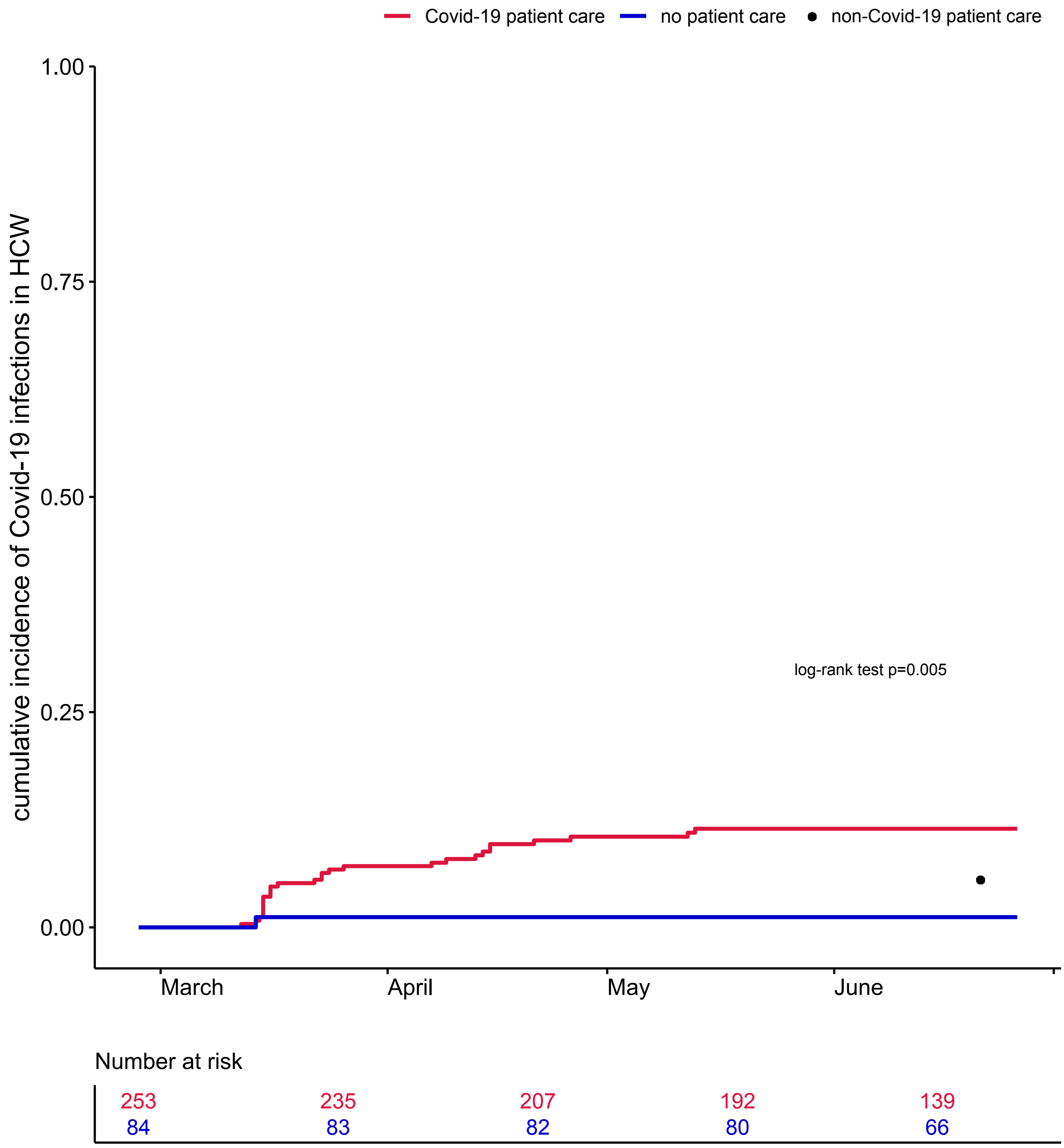

C: Cumulative incidence of SARS-CoV-2 infection in health care workers with different levels of Covid-19 patient exposure based only on nucleic acid amplification test results

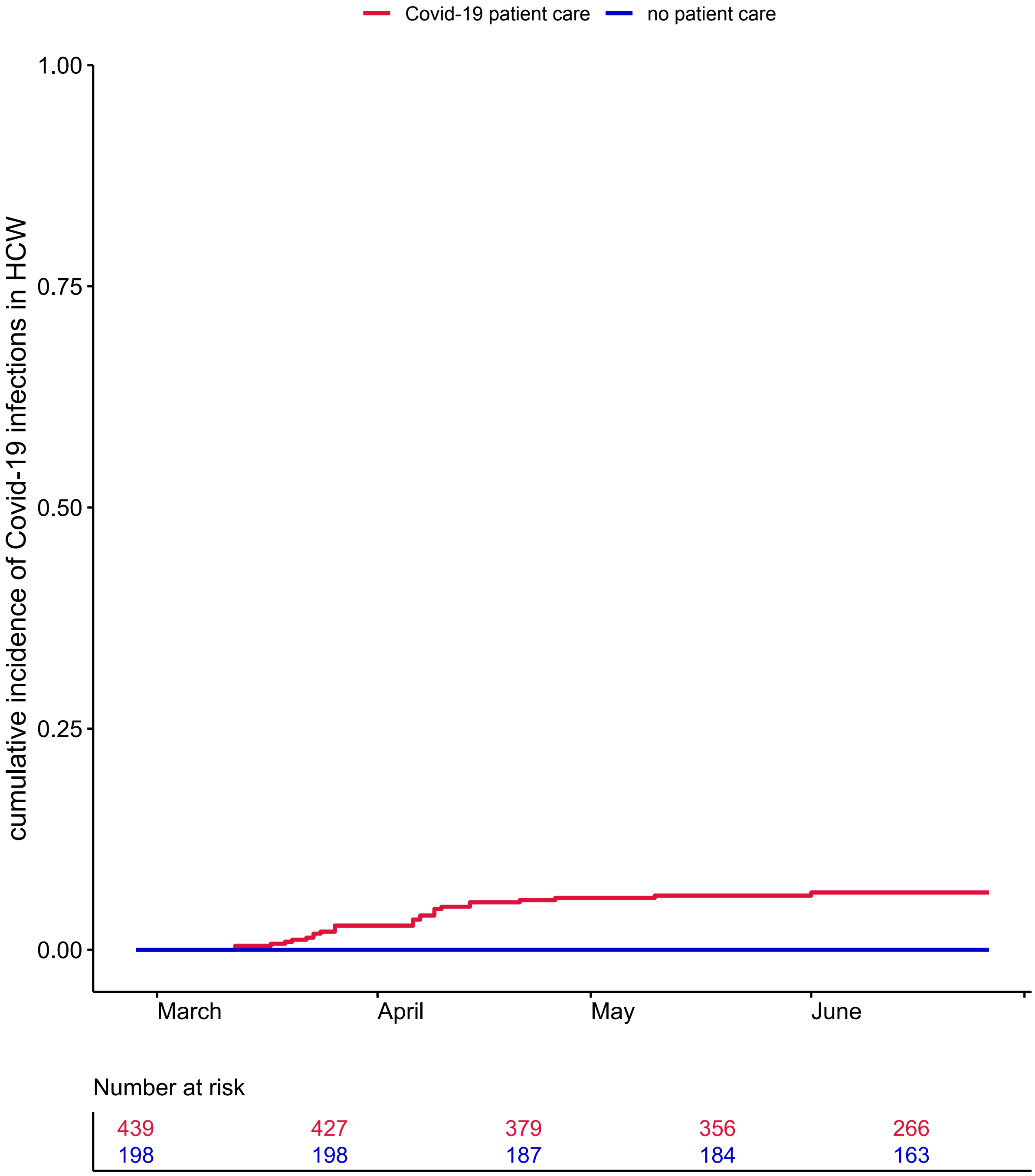

D: Cumulative incidence of SARS-CoV-2 infection in health care workers with different levels of Covid-19 patient exposure based only on serologic test results

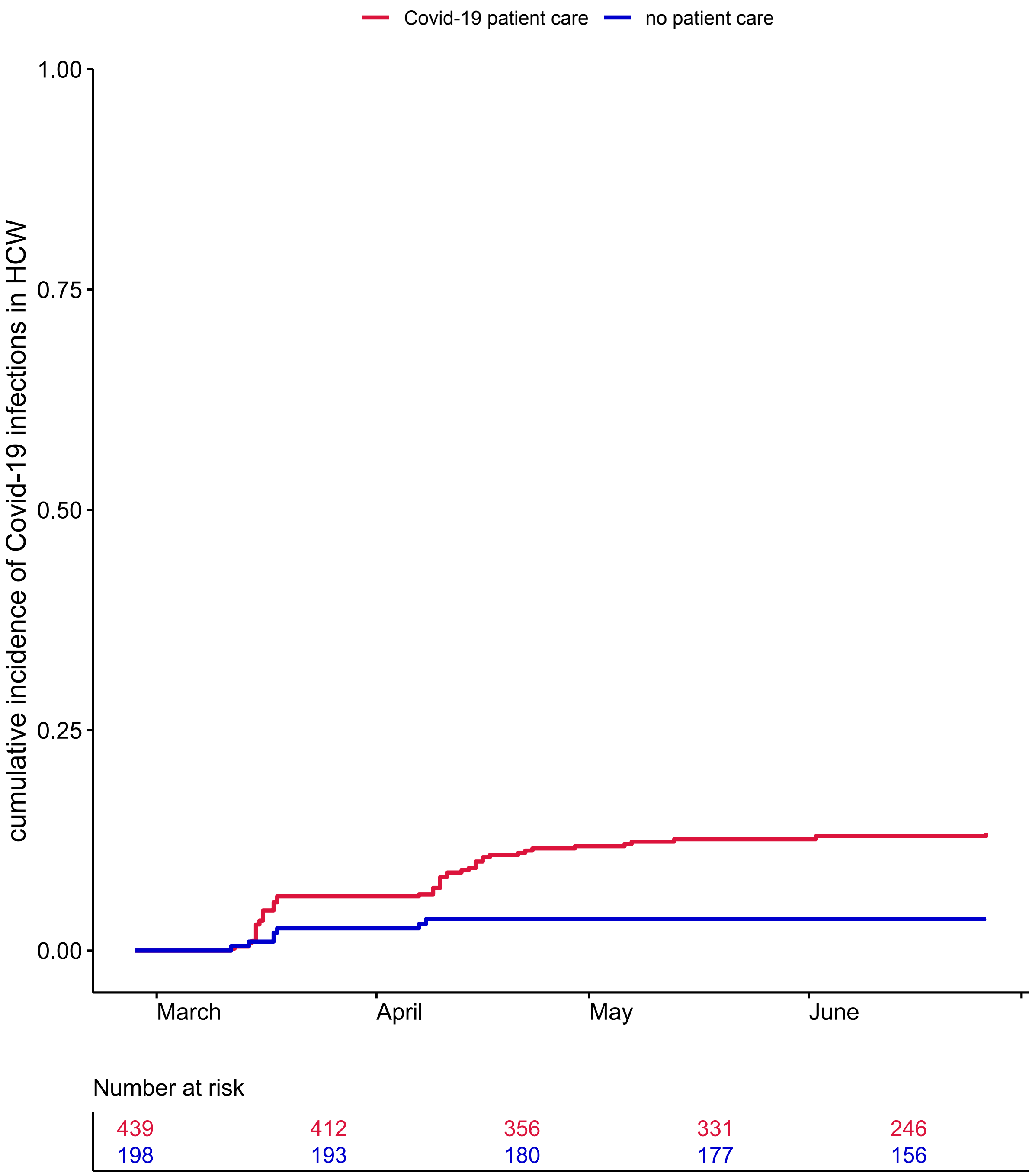

*Caption:* All participants were assumed to be seronegative on February 27 which was 4 weeks before the first measurement and the day the first Covid-19 patient was diagnosed in the Netherlands. Log-rank test reflects differences between health care workers (HCW) in Covid-19 care and HCW not in patient care (no patient care). A+B: Date of SARS-CoV-2 infection was defined as the sampling date of a first positive nucleic acid amplification test result or, in its absence, the midpoint in time between the last seronegative and the first seropositive sample. HCW in non-Covid-19 patient care (black dot) were included in the fourth measurement only and added to the figures for reference. C+D: Date of SARS-CoV-2 infection was defined as the sampling date of a first positive nucleic acid amplification test result (C) or the midpoint in time between the last seronegative and the first seropositive sample (D).

**Figure S2**

A: Cumulative incidence of SARS-CoV-2 infection in health care workers defined by hospital unit type for hospital 1

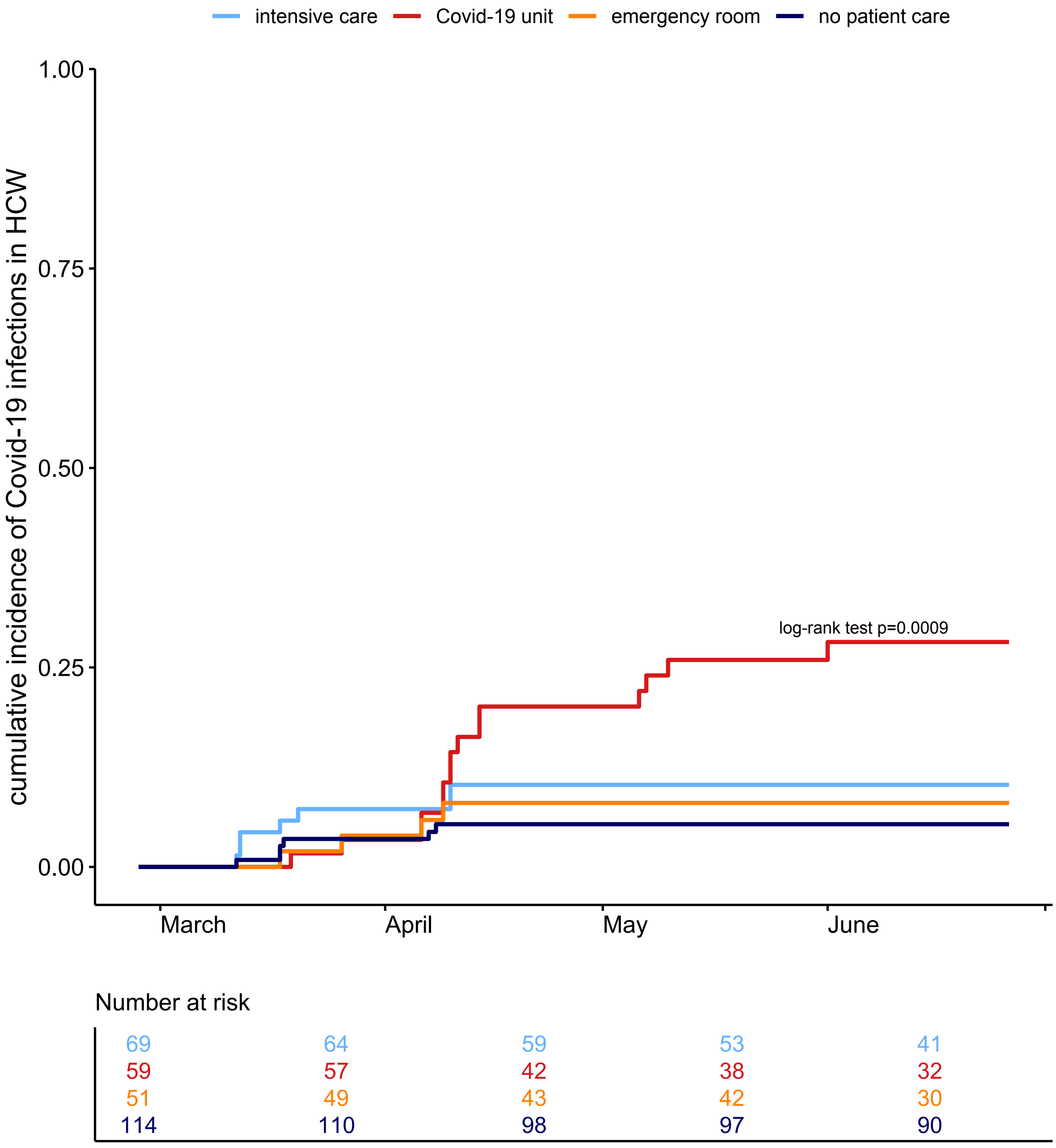

B: Cumulative incidence of SARS-CoV-2 infection in health care workers defined by hospital unit type for hospital 2

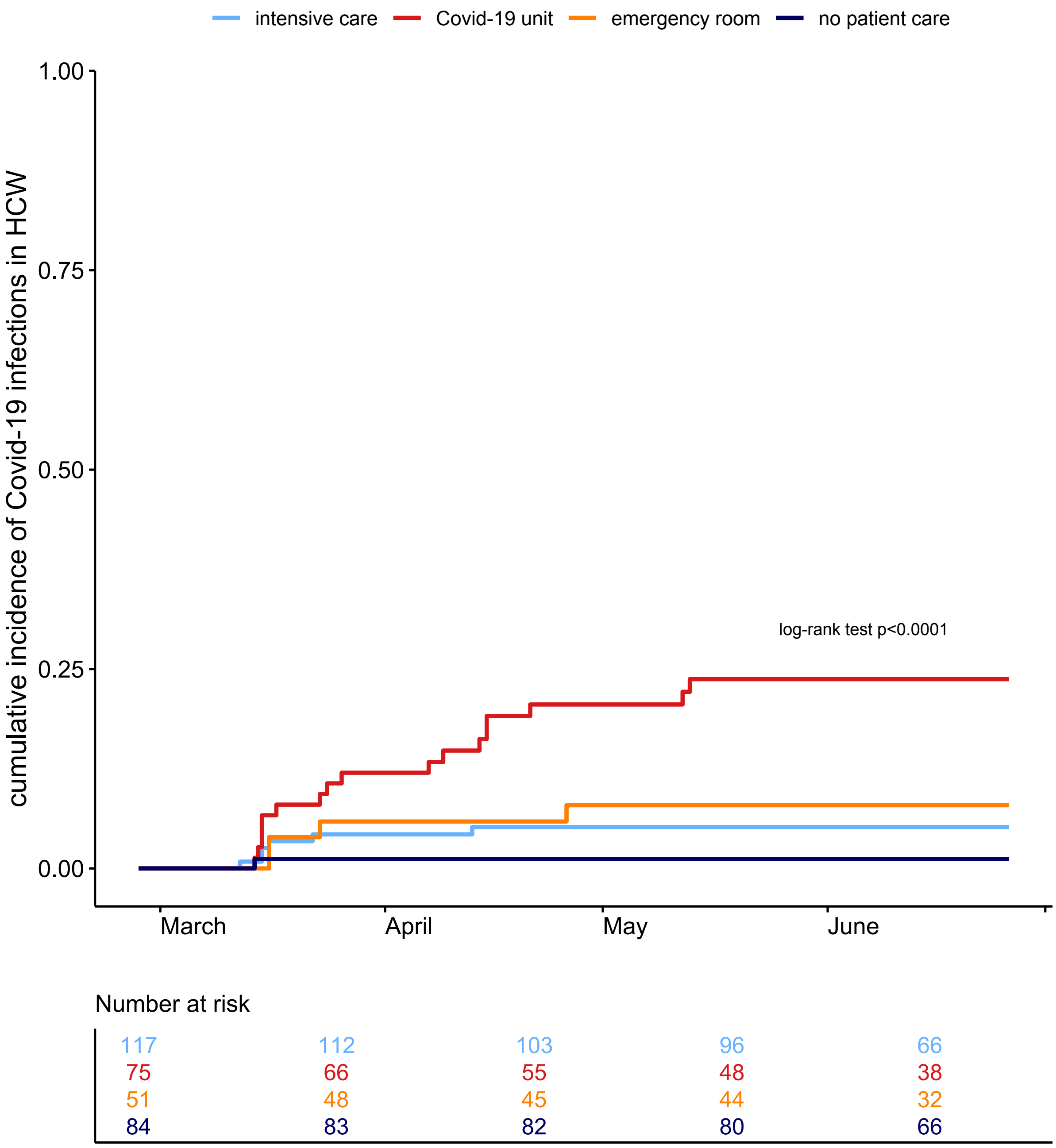
intensive care
Covid-19 unit
emergency room
no patient care
cumulative incidence of Covid-19 infections in HCW
1.00
0.75
0.50
0.25
0.00
log-rank test p<0.0001
March
April
May
June
Number at risk
117 112 103 96 66
75 66 55 48 38
51 48 45 44 32
84 83 82 80 66

C: Cumulative incidence of SARS-CoV-2 infection in health care workers defined by hospital unit type based only on nucleic acid amplification test results

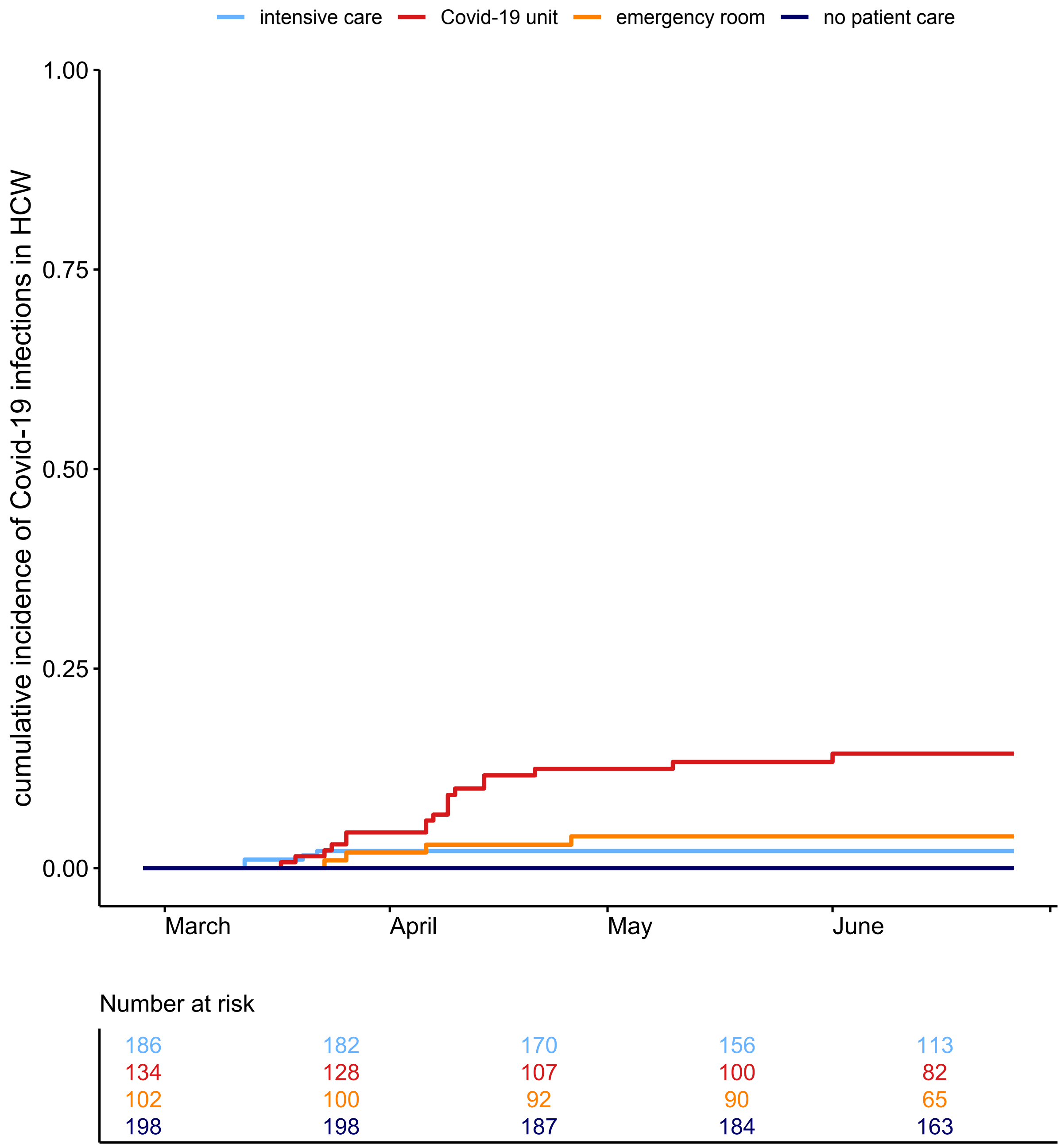

D: Cumulative incidence of SARS-CoV-2 infection in health care workers defined by hospital unit type based only on serologic test results

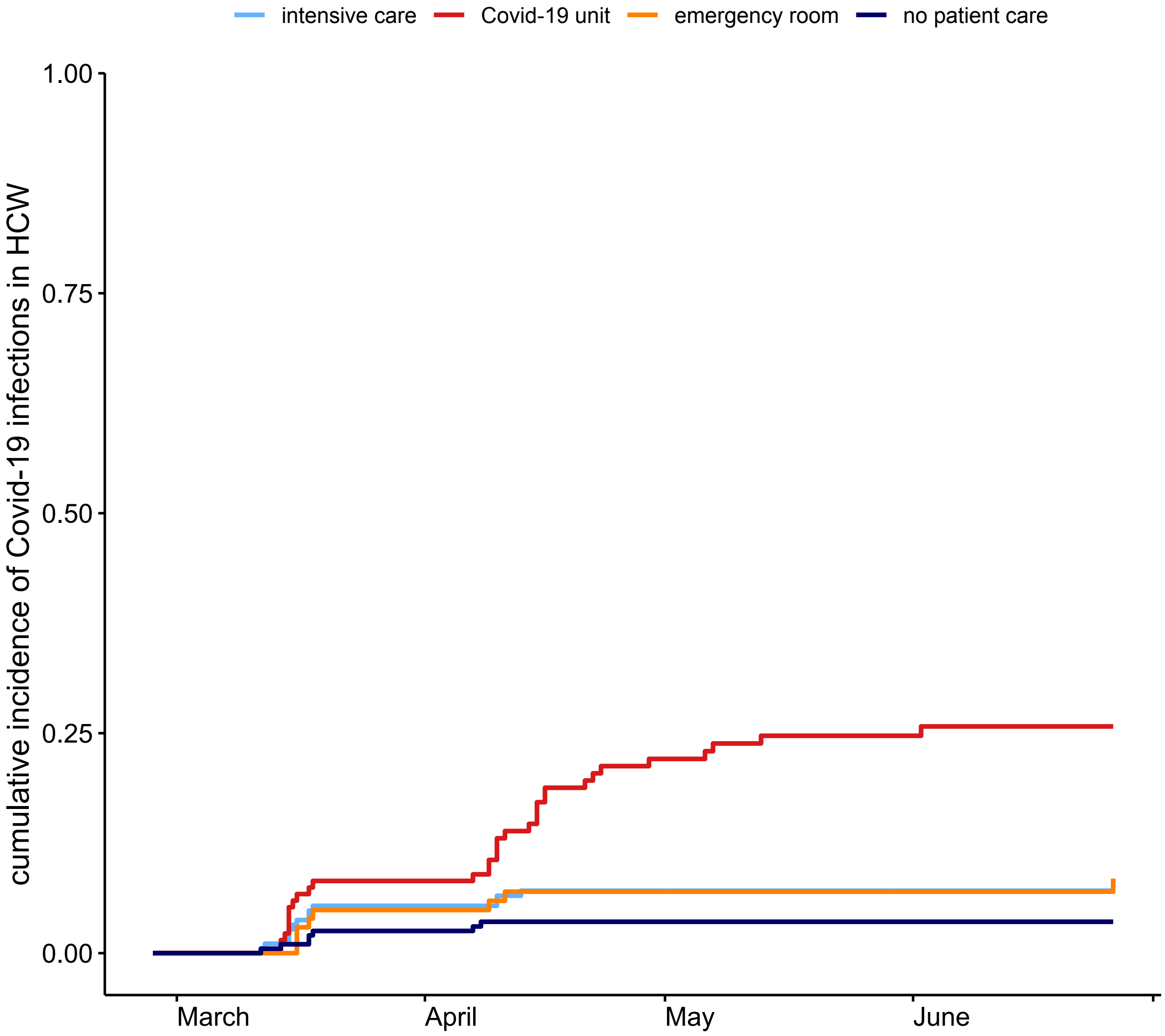

Number at risk

| | | | | |
|---|---|---|---|---|
| 186 | 176 | 162 | 149 | 107 |
| 134 | 123 | 96 | 86 | 70 |
| 102 | 97 | 89 | 87 | 63 |
| 198 | 193 | 180 | 177 | 156 |

*Caption:* Log-rank test reflects differences between all HCW groups shown. Participants working on multiple hospital unit types during the study were excluded from this analysis because of small group size. ICU, intensive care unit.

**Figure S3**

Cumulative incidence of SARS-CoV-2 infection in health care workers per specific Covid-19 regular ward

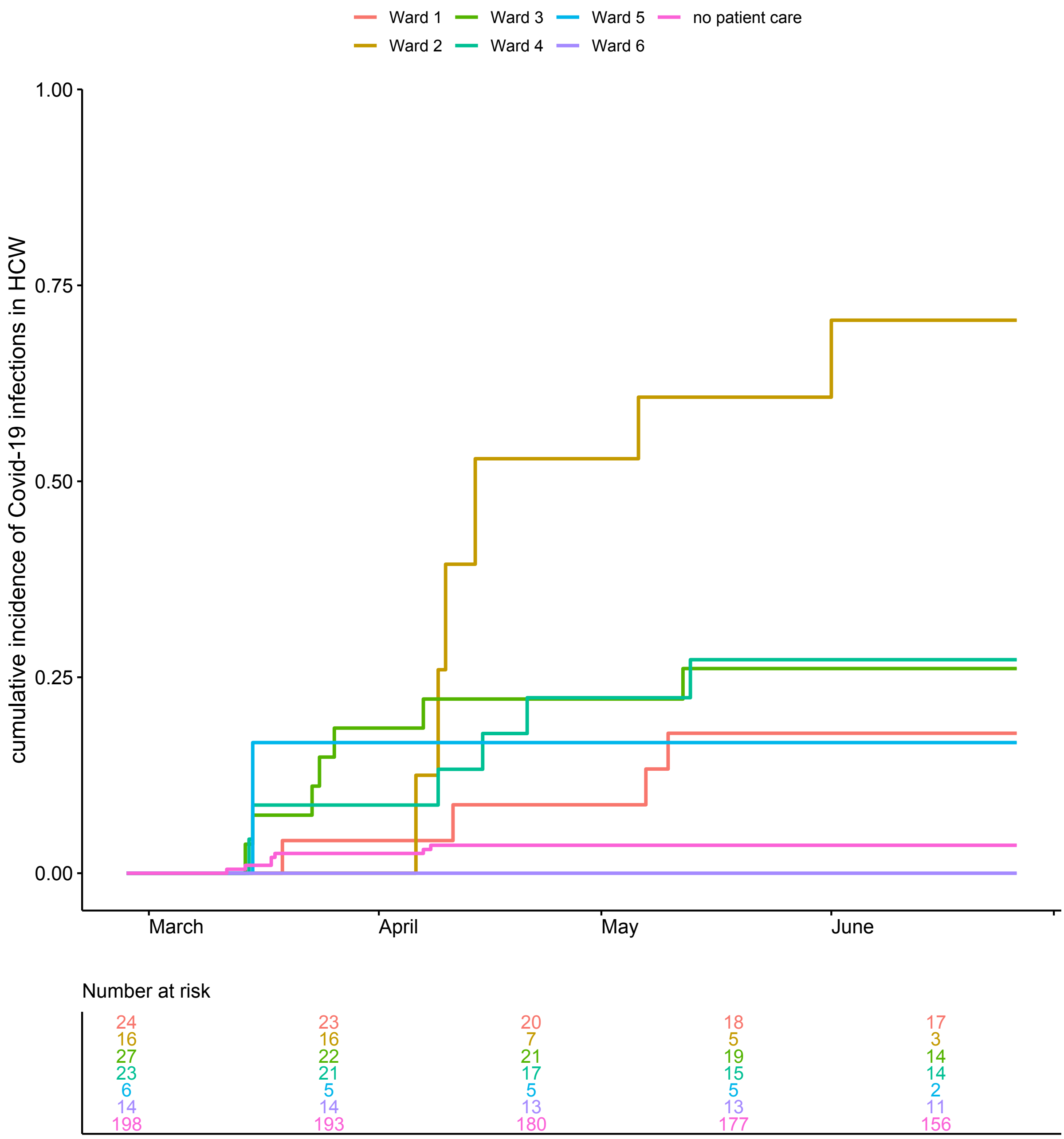

*Caption:* All participants were assumed to be seronegative on February 27 which was 4 weeks before the first measurement and the day the first Covid-19 patient was diagnosed in the Netherlands. Date of SARS-CoV-2 infection was defined as the sampling date of a first positive nucleic acid amplification test result or, in its absence, the midpoint in time between the last seronegative and the first seropositive sample.

### Figure S4

Working shifts and admittance dates of individuals identified in each transmission cluster

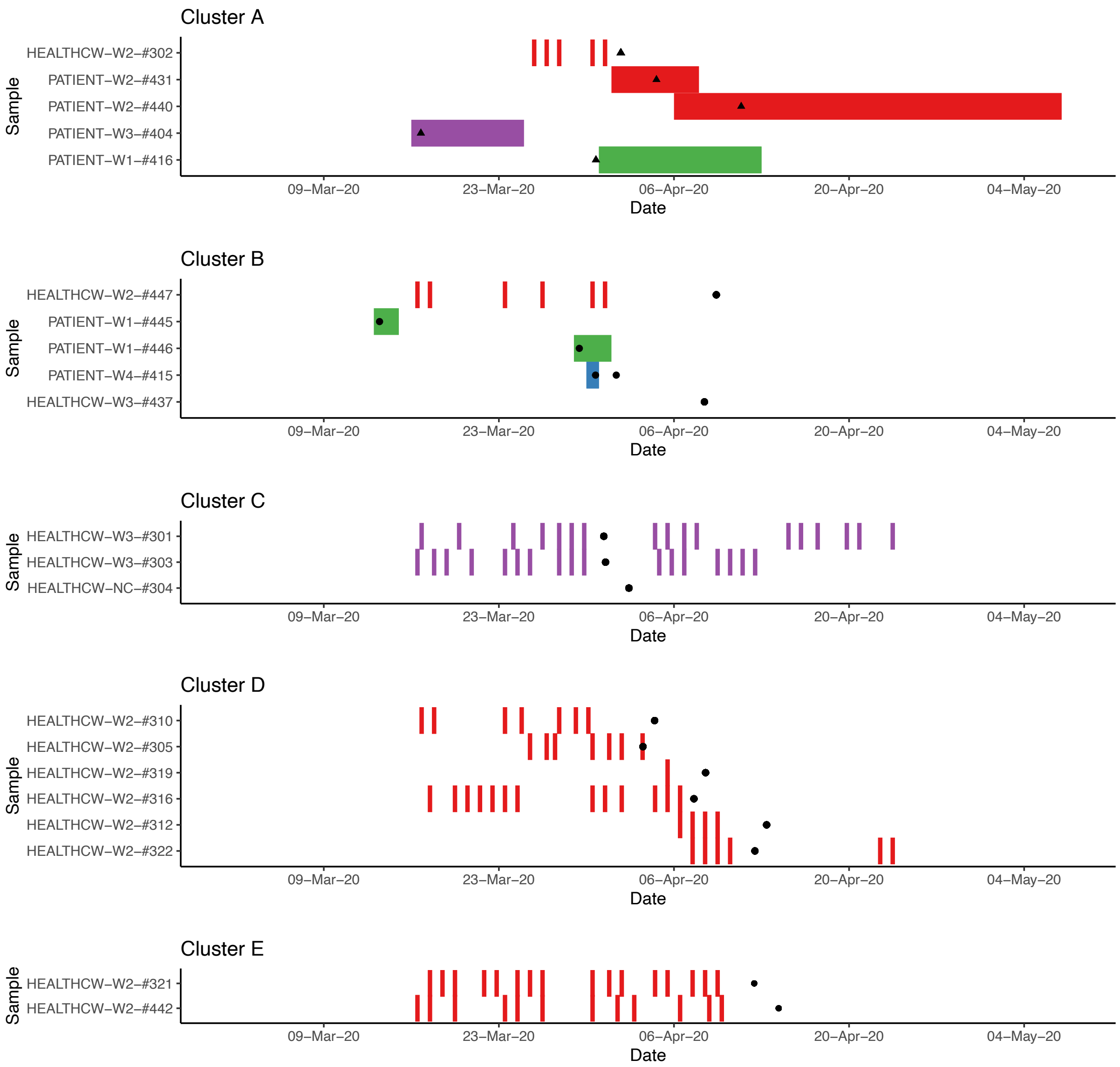

*Caption:* Panels show timeline of working shifts and admittance of health care workers and patients respectively. Black symbols (triangles for health care workers, circles for patient) denote the date of positive nucleic acid amplification test. The missing timelines in Cluster B and C are due to unknown shift schedules of the HCW. There was a lack of clear epidemiological links between patients in clusters A and B i.e., no overlapping admission dates) with at least 1 HCW working shift or patient admission at the same ward while two patients in cluster A did have overlapping admission times on the same ward.

### Supplemental table

#### Table S1

Results of univariable and multivariable cox regression analysis of association between SARS-CoV-2 infection and determinants in the overall study population (A) and within Covid-19 patient care (B) after exclusion of ward 2

| **A: univariable and multivariable cox regression models** | | | | |
|---|---|---|---|---|
| | | SARS-CoV-2 incidence N/total (%, 95% CI)) | HR (95% CI) | Adjusted HR (95% CI) |
| HCW work environment | no patient care | 7/198 (3.6, 0.9-6.1) | 1 | 1 |
| | non-COVID-19 patient care only | 11/164 (6.7, 2.8-10.5) | 1.8 (0.7-4.5) | 1.5 (0.6-4.2) |
| | COVID-19 patient care | 44/423 (11.0, 7.9-14.0) | 3.3 (1.5-7.3) | 2.4 (0.9-6.0) |
| COVID-19 coworker contact | no | 28/452 (6.6, 4.2-8.9) | 1 | 1 |
| | yes | 32/306 (11.3, 7.6-14.9) | 1.8 (1.1-3.0) | 1.6 (0.86-2.8) |
| COVID-19 community contact | no | 44/680 (7.1, 5.0-9.0) | 1 | 1 |
| | yes | 18/105 (18.7, 10.4-26.3) | 2.8 (1.6-4.8) | 2.4 (1.3-4.3) |
| days/week spent in hospital | - | - | 1.1 (0.8-1.4) | 1.6 (0.86-2.8) |
| age | - | - | 0.98 (0.96-0.998) | 0.99 (0.96-1.01) |
| **B: univariable and multivariable cox regression models within COVID-19 patient care group** | | | | |
| | | SARS-CoV-2 incidence N/total (%, 95% CI) | HR (95% CI) | Adjusted HR (95% CI) |
| hospital unit type | intensive care | 13/186 (7.1, 3.3-10.7) | 1 | 1 |
| | COVID-19 unit | 22/118 (19.7, 12.0-26.8) | 2.8 (1.4-5.5) | 2.8 (1.2-6.7) |

| | | | | |
|---|---|---|---|---|
| | emergency room | 7/102 (8.0, 2.5-13.1) | 1.1 (0.5-2.7) | 1.2 (0.4-3.4) |
| | combination of above | 2/17 (11.8, 0.0-25.8) | 2.0 (0.4-8.7) | 1.3 (0.1-13.4) |
| position | specialist | 4/84 (4.8, 0.1-9.3) | 1 | 1 |
| | resident | 12/104 (12.8, 6.0-19.1) | 2.9 (0.94-8.8) | 1.8 (0.5-6.8) |
| | nurse | 28/235 (12.4, 8.0-16.6) | 2.6 (0.91-7.4) | 1.5 (0.5-4.9) |
| self-reported COVID-19 exposure | low | 11/86 (13.2, 5.6-20.2) | 1 | 1 |
| | medium | 13/160 (9.2, 4.5-13.7) | 0.7 (0.3-1.4) | 0.5 (0.2-1.3) |
| | high | 11/68 (16.4, 7.1-24.9) | 1.2 (0.5-2.7) | 1.2 (0.4-3.5) |
| | very high | 9/109 (8.4, 3.0-13.5) | 0.6 (0.2-1.4) | 0.7 (0.2-2.2) |
| feasibility social distancing | easy | 0/2 (0.0, 0.0-0.0) | NA | NA |
| | medium | 5/25 (20.0, 2.7-34.2) | 1 | 1 |
| | difficult | 13/101 (13.4, 6.3-19.9) | 0.6 (0.2-1.7) | 0.4 (0.1-1.5) |
| | virtually impossible | 24/274 (9.3, 5.8-12.7) | 0.4 (0.2-1.1) | 0.4 (0.1-1.2) |
| PPE always correctly used | no | 5/53 (9.5, 1.2-17.1) | 1 | 1 |
| | yes | 39/368 (11.3, 7.9-14.6) | 1.2 (0.5-2.9) | 1.0 (0.3-3.0) |
| COVID-19 coworker contact | no | 15/183 (8.3, 4.2-12.3) | 1 | 1 |
| | yes | 27/219 (13.0, 8.4-17.4) | 1.6 (0.84-2.9) | 2.4 (1.0-5.4) |

| | | | | |
|---|---|---|---|---|
| COVID-19 community contact | no | 31/347 (9.6, 6.4-12.7) | 1 | 1 |
| | yes | 13/76 (17.6, 8.4-25.8) | 1.9 (1.016-3.7) | 1.5 (0.7-3.3) |
| days/week spent in hospital | - | - | 0.7 (0.5-1.0) | 0.7 (0.5-1.0) |
| age, years | - | - | 0.97 (0.94-1.001) | 0.99 (0.96-1.03) |

*Caption:* HR, hazard ratio; CI, confidence interval. Percentages with confidence intervals were calculated using the Kaplan Meier method. Adjusted HRs shown in A belong to a model containing all variables in A, likewise for B.
